# Pretreatment Identification of 90-Day Readmission Among Heart Failure Patients Receiving Aquapheresis Treatment

**DOI:** 10.1101/2023.05.05.23289603

**Authors:** Deya Alkhatib, Ibrahim Karabayir, Issa Pour-Ghaz, Sania Khan, Linda Hart, Megan Cease, John L Jefferies, Oguz Akbilgic

**Affiliations:** The University of Tennessee Health Science Center – Division of Cardiovascular Disease, Memphis, TN, 38163, USA; Wake Forest School of Medicine – Section of Cardiovascular Medicine, Winston-Salem, NC, 27157, USA; Senior Clinical Education Manager – Nuwellis, Eden Prairie, MN, 55344, USA; Vice President of Clinical Research – Nuwellis, Plymouth, MN, 55446, USA

**Keywords:** AI, Artificial Intelligence, Aquapheresis, Heart failure, HF, Hospitalization, Machine learning, ML, Predicting heart failure, Readmission, Ultrafiltration

## Abstract

**BACKGROUND:** The use of traditional models to predict heart failure (HF) has limitations in preventing HF hospitalizations. Artificial intelligence (AI) and machine learning (ML) in cardiovascular medicine only have limited data published regarding HF populations, with none assessing the favorability of decongestive therapy aquapheresis (AQ). AI and ML can be leveraged to design non-traditional models to identify those who are at high risk of HF readmissions.

**OBJECTIVES:** This study aimed to develop a model for pretreatment identification of risk for 90-day HF events among HF patients who have undergone AQ.

**METHODS:** Using data from the AVOID-HF (Aquapheresis versus Intravenous Diuretics and Hospitalization for Heart Failure) trial, we designed a ML-based predictive model that can be used before initiating AQ to anticipate who will respond well to AQ and who will be at high risk of future HF events.

**RESULTS:** Using ML we identified the top ten predictors for 90-day HF events. Interestingly, the variable for ‘intimate relationships with loved ones’ strongly predicted response to therapy. This ML-model was more successful in predicting the outcome in HF patients who were treated with AQ. In the original AVOID-HF trial, the overall 90-day HF event rate in the AQ arm was 32%. Our proposed predictive model was accurate in anticipating 90-day HF events with better statistical accuracy (area under curve 0.88, sensitivity 80%, specificity 75%, negative predictive value 90%, and positive predictive value 57%).

**CONCLUSIONS:** ML can help identify HF patients who will respond to AQ therapy. Our model can identify super-respondents to AQ therapy and predict 90-day HF events better than currently existing traditional models.

**CONDENSED ABSTRACT:** Utilizing data from the AVOID-HF trial, we designed a ML-predictive model that can be used before initiating AQ to anticipate who will respond well to AQ and who will be at high risk of future HF events. Using ML, we identified the top 10 predictors for 90-day HF events. Our model can identify super-respondents to ultrafiltration therapy and predict 90-day HF events better than currently existing traditional models.

## 1. Introduction

Heart failure (HF) is a very challenging and complex clinical syndrome due in part to the heterogeneity of patient phenotype. HF prevalence continues to rise along with population aging. Approximately 6.2 million American adults had HF from 2013 to 2016 compared to 5.7 million patients between 2009 and 2012^1^. More than eight million patients are estimated to live with HF by 2030^2^. HF phenotypes have complex heterogeneity due to their different clinical characteristics, responses to treatment, and outcomes. These factors often contribute to frequent hospitalization with documented high readmission rates. HF results in a significant burden on communities and healthcare systems.

Fluid accumulation initially shows no manifestations before the intravascular volume is expanded and congestion manifests. Congestion in HF patients is a leading cause of emergency department (ED) visits and accounts for most HF hospitalizations^3^. Nearly half of HF patients are discharged with unresolved congestion setting them up for recurrent rehospitalizations^4^. Guidelines recommend optimizing volume status in HF patients with fluid retention to improve their symptoms and achieve euvolemic status^5^. Loop diuretics (LD) work by blocking the Na/K/2Cl co-transporter in the ascending loop of Henle and creating an osmotic gradient essential for water reabsorption resulting in natriuresis. Low urine sodium (Na) concentration or reduced response to diuretics, known as restricted diuretic response or diuretic resistance (DR), can cause renal tubular injury. Diuretics can activate the unwanted neurohormonal sympathetic system including the renin-angiotensin-aldosterone system (RAAS) which can explain DR^6^. Distal tubular Na reabsorption is another cause of DR. In order to overcome DR, current guidelines recommend escalating LD dose, changing oral to intravenous (IV) diuretics, or combining different diuretics classes (thiazide or thiazide-like diuretic)^7^. In a propensity-adjusted cohort study among hospitalized patients with acute decompensated heart failure (ADHF), the addition of a thiazide-like diuretic (metolazone) independently increased the risk of hyponatremia, hypokalemia, worsening kidney function, and mortality rate. On the other hand, the use of high doses of LD was not associated with reduced survival after propensity adjustment. However, there are limited randomized controlled trials comparing those strategies^8^.

Ultrafiltration (UF) is an alternative, effective, and safe method of removing Na and water in HF patients. The RAPID-CHF (the Relief for Acutely Fluid-Overloaded Patients With Decompensated Congestive Heart Failure) trial assessed the safety and efficacy of UF in patients admitted with ADHF. It showed that early UF in ADHF patients was safe and well-tolerated^9^. The EUPHORIA (early ultrafiltration therapy in patients with decompensated heart failure and observed resistance to intervention with diuretic agents) trial showed that in patients with ADHF and DR, UF before IV diuretics was safe and effective in shortening the length of stay and decreasing the readmission rate^10^. The UNLOAD (Ultrafiltration versus Intravenous Diuretics for Patients Hospitalized for Acute Decompensated Congestive Heart Failure) trial compared UF to IV diuretics for patients hospitalized with ADHF. It showed that UF was more effective in weight and fluid loss and reducing 90-day readmission rate compared to IV diuretics. With the safety and positive primary and secondary outcome results, UF appeared to be an effective and alternative therapy for ADHF patients^11^. The ULTRADISCO (ULTRAfiltration vs. DIureticS in deCOmpensated HF) trial evaluated the hemodynamic and clinical effects of UF versus IV diuretics in ADHF patients. It showed that ADHF patients treated with UF had a better clinical improvement and a significant reduction in plasma aldosterone level and N-terminal (NT)-pro hormone BNP (NT-proBNP). Systemic vascular resistance index was significantly reduced 36 hours after treatment with UF compared to IV diuretics. This study was able to show that UF appears to overcome the pathophysiologic issues seen due to unwanted RAAS and neurohormonal activation associated with diuretics use^12^.

The CARRESS-HF (Cardiorenal Rescue Study in Acute Decompensated Heart Failure) trial assessed the safety and efficacy of UF in ADHF patients complicated by worsened renal function and persistent congestion. This randomized trial compared UF to stepped pharmacologic therapy assessing the primary outcome of serum creatinine level and body weight 96 hours after randomization. It showed that stepped pharmacologic therapy was superior to UF strategy for renal function preservation (−0.04±0.53 mg per deciliter versus +0.23±0.70 mg per deciliter; P=0.003) with no difference in weight loss between the two groups. A higher incidence of serious adverse events was seen in the UF group (72% versus 57%; P=0.03) ^13^. This important trial with negative results raised many questions and concerns about UF safety and effectiveness. The CARRESS-HF trial had many limitations, including the methodology and protocol. For instance, more than one-third of the UF group received diuretics instead of UF or received diuretics after stopping UF prior to intention to treat analysis, which questioned the accurate assessment of adverse events and renal function in both groups. Another limitation was that the UF rate was fixed (200 mL/hour) for all patients in the UF group, whereas stepped pharmacologic therapy was titrated based on urine output (goal to achieve 3-5 L of urine/day). Also, 12% of patients in the pharmacologic therapy group received inotropic agents that were prohibited in the UF arm. At the time of assessment and comparison (96 hours after randomization), only one-third of the UF group were still included in the study protocol compared to 80% of the pharmacologic therapy group^14^. The CUORO (continuous ultrafiltration for congestive heart failure) trial, a small, randomized, single-center study, compared UF (adjustable UF rate) to standard medical therapy in ADHF patients with a primary endpoint of HF hospitalization rate at one year. A lower rate of HF hospitalization in one year was seen in the UF group (hazard ratio 0.14, 95% confidence interval 0.04-0.48; P = 0.002)^15^. With the positive result of the CUORO trial along with the UNLOAD trial results, it appeared that UF has a prolonged and protective effect. Results from the CARRESS-HF and CUORO trials encouraged researchers to design some larger, randomized, and multicenter trials.

The AVOID-HF (Aquapheresis versus Intravenous Diuretics and Hospitalization for Heart Failure) trial, a randomized, large, multicenter study, was designed to assess whether adjustable UF can prolong the time to first HF event within 90 days of hospital discharge, compared to adjustable LD therapy. The fluid removal strategy in both arms was adjustable based on vital signs and kidney function. Among patients who were hospitalized for ADHF, a total of 110 patients were randomized to adjustable UF and 114 patients were randomized to adjustable LD. UF was performed using the Aquadex FlexFlow System (Baxter International, Deerfield, Illinois). Results showed that the adjustable UF arm had more days to first HF events (62 days versus 34 days for the adjustable LD arm; P = 0.106) and fewer HF events (25% vs. 35%; hazard ratio of 0.663; 95% confidence interval: 0.402 to 1.092); HF events were defined as either unscheduled outpatient or ED visits for ADHF treatment with IV LD or UF, or HF rehospitalization ^16^. The 90-day mortality rate was similar, however, the adjustable UF arm had more adverse effects of special interest (P = 0.018) and serious adverse events related to the study product (P = 0.026). In addition, the UF arm had a greater net fluid loss^17^. The encouraging results of the AVOID-HF and UNLOAD trials showed that early UF is effective and safe with prolonged effects when used before developing kidney dysfunction^18^. A per-protocol analysis on the CARRESS-HF trial showed that UF patients, who actually received UF per study protocol, had significantly more weight and fluid loss; this was accompanied by increased serum creatinine and neurohormonal activation in the UF group. These elevations in biomarkers correlated with superior decongestion and renal function recovery at 60 days ^19^.

## 2. Methods

The AVOID-HF trial, a randomized, large, multicenter study, was designed to assess whether adjustable UF can prolong the time to the first HF event within 90 days of hospital discharge, compared to adjustable LD therapy. UF was performed using the Aquadex FlexFlow System (Baxter International, Deerfield, Illinois).

### 2.1. Objective and Outcome

The purpose of the study was to develop a model for pretreatment identification of risk for 90-day HF event among HF patients who have undergone AQ

### 2.2. Study Population

Patients with missing 90-day follow-up, including those who died within 90 days of treatment, were excluded.

### 2.3. Cross-validation Setup

To avoid overfitting, we implemented a two-level cross-validation setup. To do that, we first split the analytical cohort into training and hold-out sets at 80:20 proportion. We then carried out a leave-one-out cross-validation on the 80% training data. The final cross-validated model was further implemented on the 20% untouched hold-out data.

### 2.4. Design of the machine-learning algorithm for predicting 90-day events

We utilized the Light Gradient Boosting Machine (LightGBM) algorithm to predict the risk for 90-day HF events. LightGBM algorithm is a decision-tree-based algorithm with several hyperparameters that need to be tuned to have a generalizable and robust model^20^. We have utilized the Bayesian Optimization approach for hyperparameters tuning during leave-one-out cross-validation on training data. For comparison, we repeated our analysis using other ML methods, including extreme gradient boosting machines, support vector machines, classification and regression trees, and random forest.

### 2.5. Variable Selection and Direction Analysis

We implemented a previously developed Feature Importance and Direction Analysis (FIDA)^21^ to identify the most important predictors of 90-day readmission and identify the direction of the interaction between input and output. In FIDA, the importance of a predictor is assessed by artificially increasing its value by one standard deviation (or swapping with a reference category for nominal variables or increasing one unit for ordinal variables) and assessing the change in predicted risk compared. FIDA is applied on a final predictive model for each predictor separately. We used FIDA to identify the most important predictors to build more compact models and better interpret our ML results.

## 3. Results

After excluding patients missing 90-day follow-up, including those who died within 90 days of treatment, 81 patients in the AQ group were used for model development (n=81). The data was split into two sets: 80% for model building (n=64; 21 cases and 43 controls) and 20% for internal validation (n=17; 5 cases and 12 controls). Data from the IV LD arm was used for sensitivity analysis of the model. LightGBM yielded superior performance, therefore, we used LightGBM ML algorithm in our analysis.

We divided the dataset into two subsets in a stratified manner: 80% for cross-validation (n=64 including 21 with and 43 without 90-day event), and 20% for holdout test set (n=17, including 5 with and 12 without 90-day event). Since there were 21 patients with a 90-day event in the 80% training set, we implemented a leave-one-out cross-validation (equivalent to 21-fold cross-validation) on the training data to avoid overfitting. At each run, 20 folds of data were used to build a prediction model and 1-fold of data was used as a validation set (**Figure 1**).

**Figure 1.**
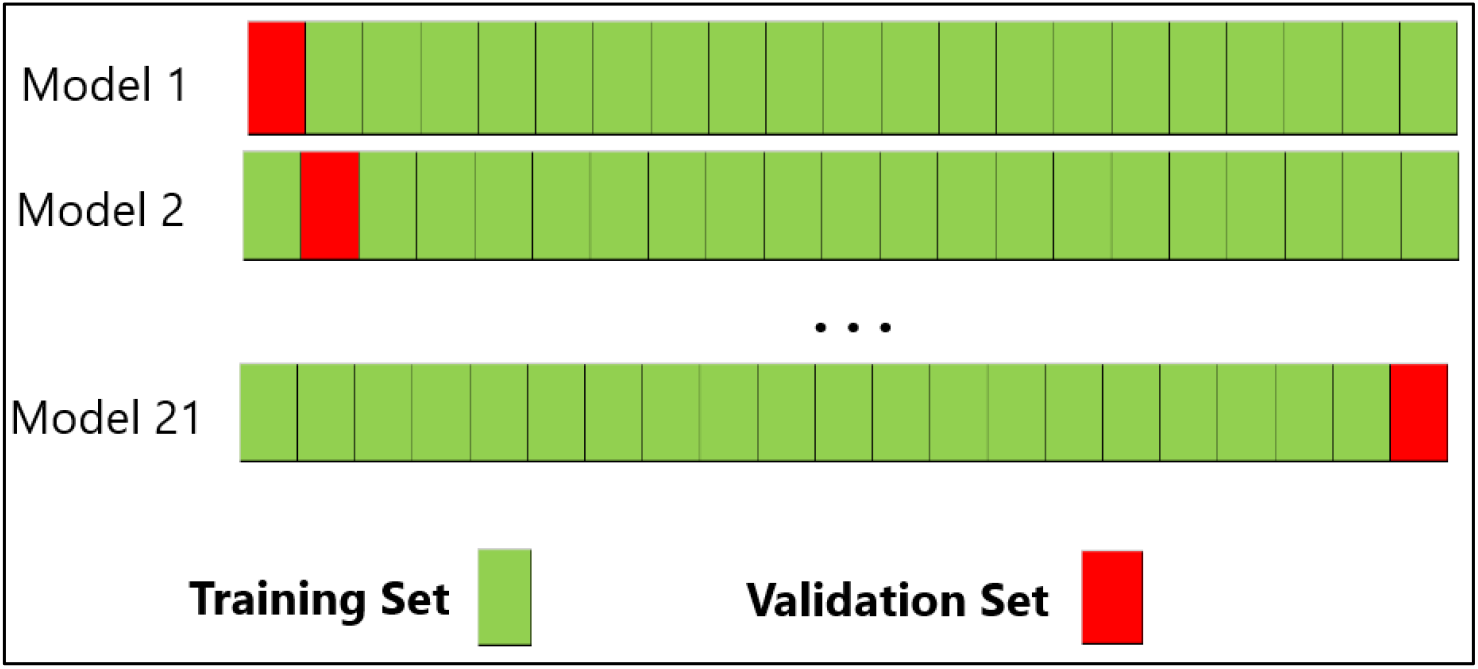
Leave-one-out cross validation for 21 instances.

This process resulted in 21 different predictive models. We then packaged these 21 models into one final ensemble model to be tested on the 20% hold-out test data. In this work, we have used the raw data with missingness to be imputed by using the default imputation algorithm of LightGBM, for both the cross-validation set and the holdout set. The Bayesian optimization algorithm found that utilizing the top 17 variables is sufficient to predict the risk for 90-day event for participants treated with Aquapheresis. We then implemented LightGBM’s default variable importance analysis. In **Table 1**, we have listed the top 17 variables in order of importance.

**Table 1.** Top 17 variables in their order of importance. ADHF: acute decompensated heart failure; IV: intravenous; LD: loop diuretic.

| Rank | Variable Name | Variable |
| --- | --- | --- |
| 1 | ACT_10_kccq_1494 | Intimate relationships with loved ones<br>• Severely limited: 0<br>• Limited quite a bit: 1<br>• Moderately limited: 2<br>• Slightly limited: 3<br>• Not limited at all: 4 |
| 2 | VALV DIS med 1494 | Valvular heart disease |
| 3 | ARRYTHST med 1494 | History of arrhythmia |
| 4 | MED NONC med 1494 | Poor adherence to medical therapy |
| 5 | DIAB med 1494 | History of diabetes mellitus |
| 6 | REF DIUR med 1494 | Suboptimal diuretic therapy response |
| 7 | COPD med 1494 | Chronic obstructive lung disease |
| 8 | DIURRCV pat 1494 | IV LD use during ADHF hospitalization |
| 9 | CVA med 1494 | History of cerebrovascular disease |
| 10 | BUMEX DIURNAM pat 1494 | IV bumetanide use |
| 11 | ICD med 1494 | Presence of implantable cardiac defibrillator |
| 12 | AORTCHST med 1494 | Aortic valve disease |
| 13 | LASIX DIURNAM pat 1494 | IV furosemide use |
| 14 | AFIBFLUT med 1494 | History of atrial fibrillation and/or atrial flutter |
| 15 | BRPACMKR med 1494 | Presence of cardiac resynchronization therapy device |
| 16 | CONCO fol 1494 | Medications change since enrollment |
| 17 | HFFACTUK med 1494 | Unknown contributing factor to worsen heart failure |

Compact Models: Unlike classic parametric regression models, additional input variables do not necessarily improve the performance. Relying on many input variables reduces the practicality of predictive models. Therefore, we further re-built models using the top 17 variables aiming to obtain a predictive ensemble model that relies on a smaller number of variables without compromising its prediction accuracy. To achieve that, we started building a new ensemble model using the top five variables from Table 1. Then, at every step, we introduced one more variable into the ensemble model based on their importance and repeated our analysis **(Figure 2)**. After adding the 10^th^ variable, the hold-out accuracy reached its maximum, and more additional variables did not improve the hold-out accuracy. The same cut-off was valid for the cross-validation set where additional variables after the 10^th^ variable reduced accuracy. Therefore, our final ensemble model is built using the top 10 input variables listed in **table 2**.

**Figure 2.**
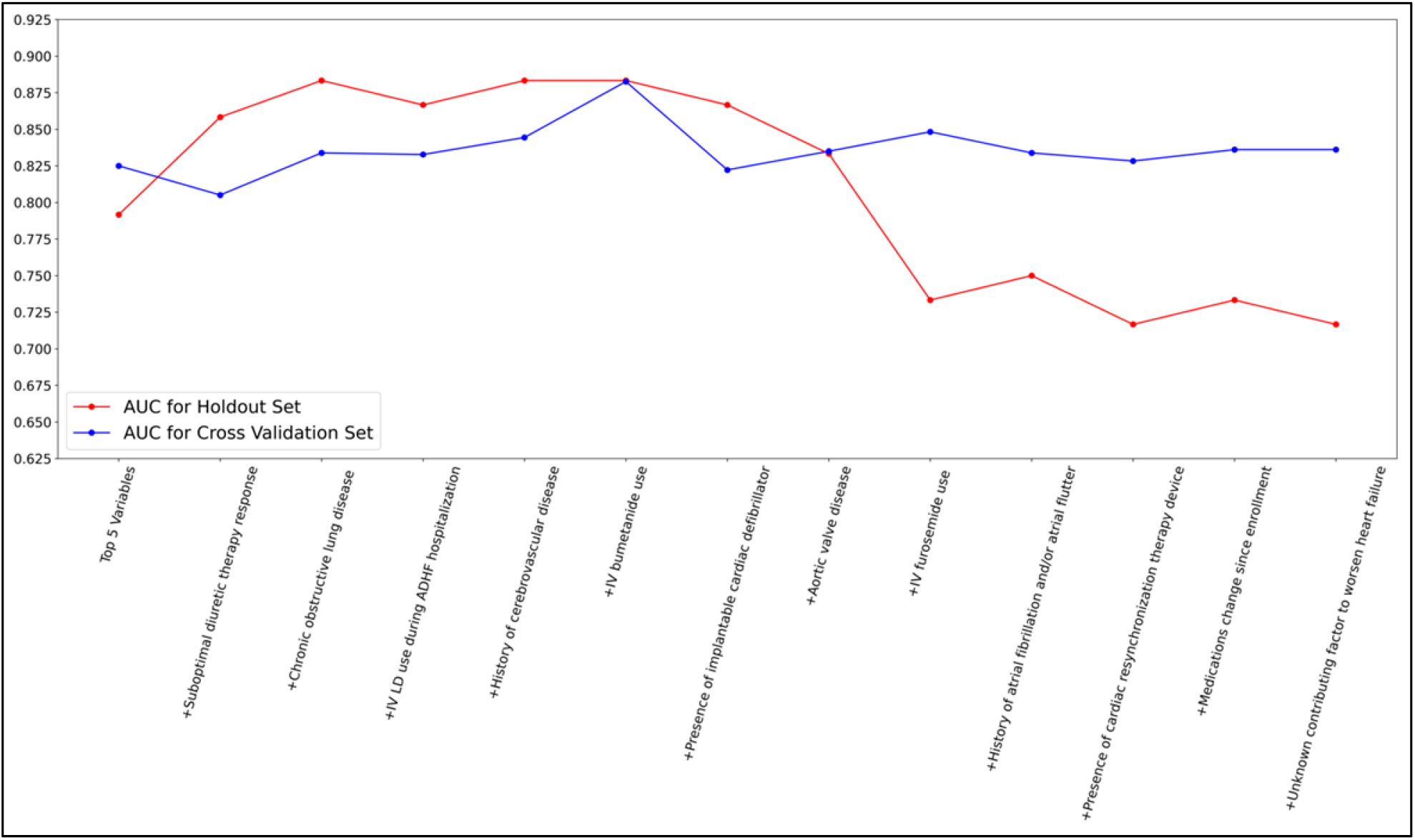
The change in the performance of the LightGBM ensemble models as new variables are introduced based on their order of importance from Table 1. Y-axis is the model performance in terms of AUC and x-axis is for input variables. ADHF: acute decompensated heart failure; AUC: area under curve; IV: intravenous; LD: loop diuretic.

**Table 2:** The top ten variables that were used in the final ensemble model. ADHF: acute decompensated heart failure; IV: intravenous; LD: loop diuretic.

| Rank | Variable |
| --- | --- |
| 1 | Intimate relationships with loved ones |
| 2 | Valvular heart disease |
| 3 | History of arrhythmia |
| 4 | Poor adherence to medical therapy |
| 5 | History of diabetes mellitus |
| 6 | Suboptimal diuretic therapy response |
| 7 | Chronic obstructive lung disease |
| 8 | IV LD use during ADHF hospitalization |
| 9 | History of cerebrovascular disease |
| 10 | IV bumetanide use |

The accuracy metrics of the final ensemble model utilizing the ten variables show area under curve (AUC) of 0.88, sensitivity (SN) of 86%, and specificity (SP) of 77% in the cross-validation set. **Table 3** summarizes the accuracy metrics for the hold-out and cross-validation sets. To get binary prediction for both sets, a cut-off value of 0.328 was set in order to balance the SN and SP in the cross-validation set; the same cut-off value was applied to the holdout set without any calibration. The AUC obtained on cross-validation and hold-out data are comparable which is a positive sign for generalizability. The negative predictive value (NPV) and positive predictive value (PPV) of the hold-out set were 90% and 57%, respectively (**Table 4**). The proposed ensemble model assigned 59% of the patients into the low-risk (or super-responders) category. Among these super-responders, 90% of them did not experience any 90-day event. On the other hand, the proposed ensemble model assigned 41% of the patients into the high-risk category. Among these patients, 57% experienced a 90-day event.

**Table 3.** Accuracy Metrics of LightGBM ensemble model. AUC: area under curve; SN: sensitivity; SP: specificity.

| Accuracy Statistics | Holdout Set (20%) | Cross Validation Set (80%) |
| --- | --- | --- |
| AUC | 0.88 [0.66 - 1.00] | 0.88 [0.8 - 0.94] |
| SN | 0.80 [0.54 - 1.00] | 0.86 [0.75 - 0.97] |
| SP | 0.75 [0.47 - 1.00] | 0.77 [0.64 - 0.9] |

**Table 4.** Confusion Matrix of Holdout Set. NPV: negative predictive value; PPV: positive predictive value; SN: sensitivity; SP: specificity.

| 90-day event |  | Actual 90-day event |  |  |
| --- | --- | --- | --- | --- |
|  |  | Yes | No |  |
| Predicted 90-day Event | Yes | 4 | 3 | PPV 57% |
|  | No | 1 | 9 | NPV 90% |
|  |  | SN 80% | SP 75% |  |

Using the final ensemble model that utilizes the top ten variables, we generated the feature importance analysis (**Figure 3**) and the direction of variables in terms of their contribution to the risk (**Figure 4**). For example, for those who reported “severely limited” intimate relationships, their average predicted risk of 90-day HF event(s) was increased by 0.18 with a standard error of 0.02. **Table 5** shows the baseline characteristics of the top ten variables used in the final ensemble model and their distribution over the predicted low and high risk for a 90-day HF event.

**Figure 3.**
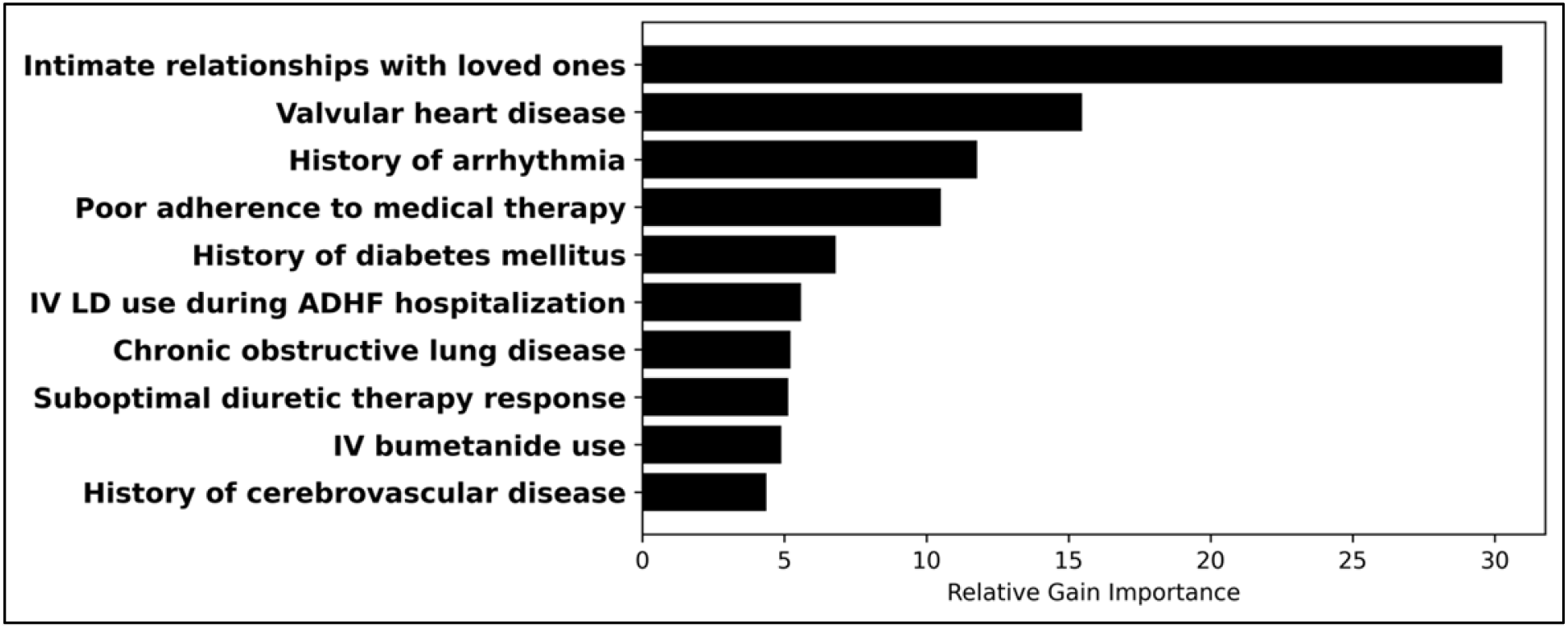
The relative contribution of the variables to the model. ADHF: acute decompensated heart failure; IV: intravenous; LD: loop diuretic.

**Figure 4:**
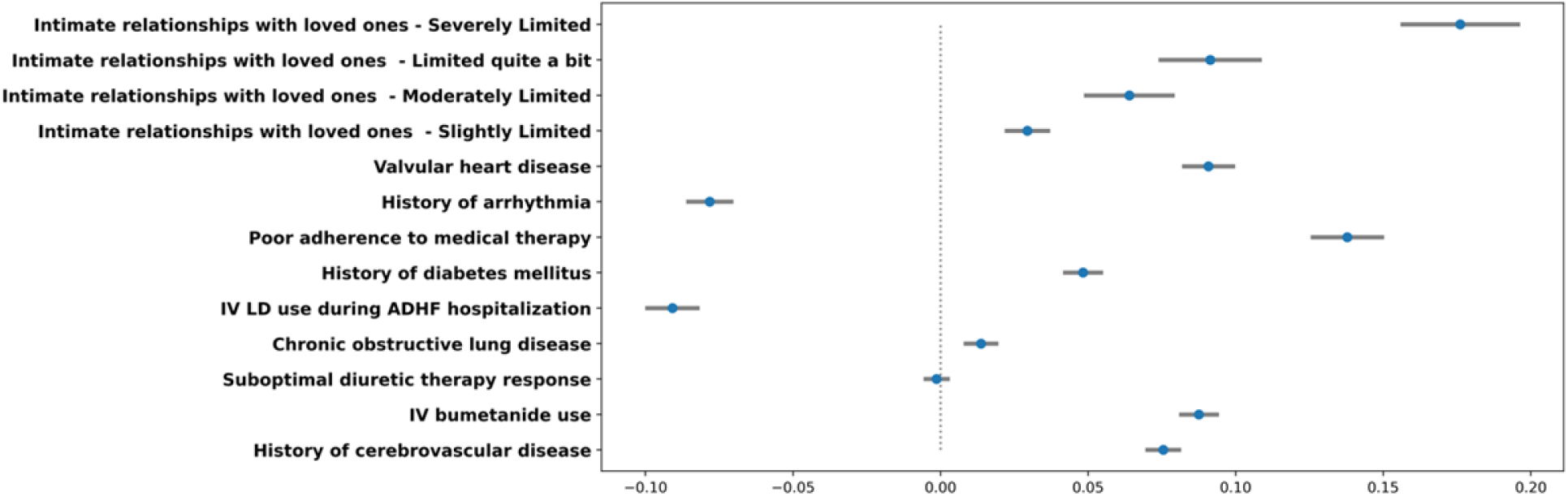
Direction of variables’ contribution to the 90-day HF event risk. Of note, the blue dot and gray line show the mean and standard error of the change in the 90-day HF event risk, respectively. For example, the 90-day readmission risk is approximately 0.18 absolute risk higher compared to patients with the same profile except with no limitation. ADHF: acute decompensated heart failure; IV: intravenous; LD: loop diuretic.

**Table 5.**
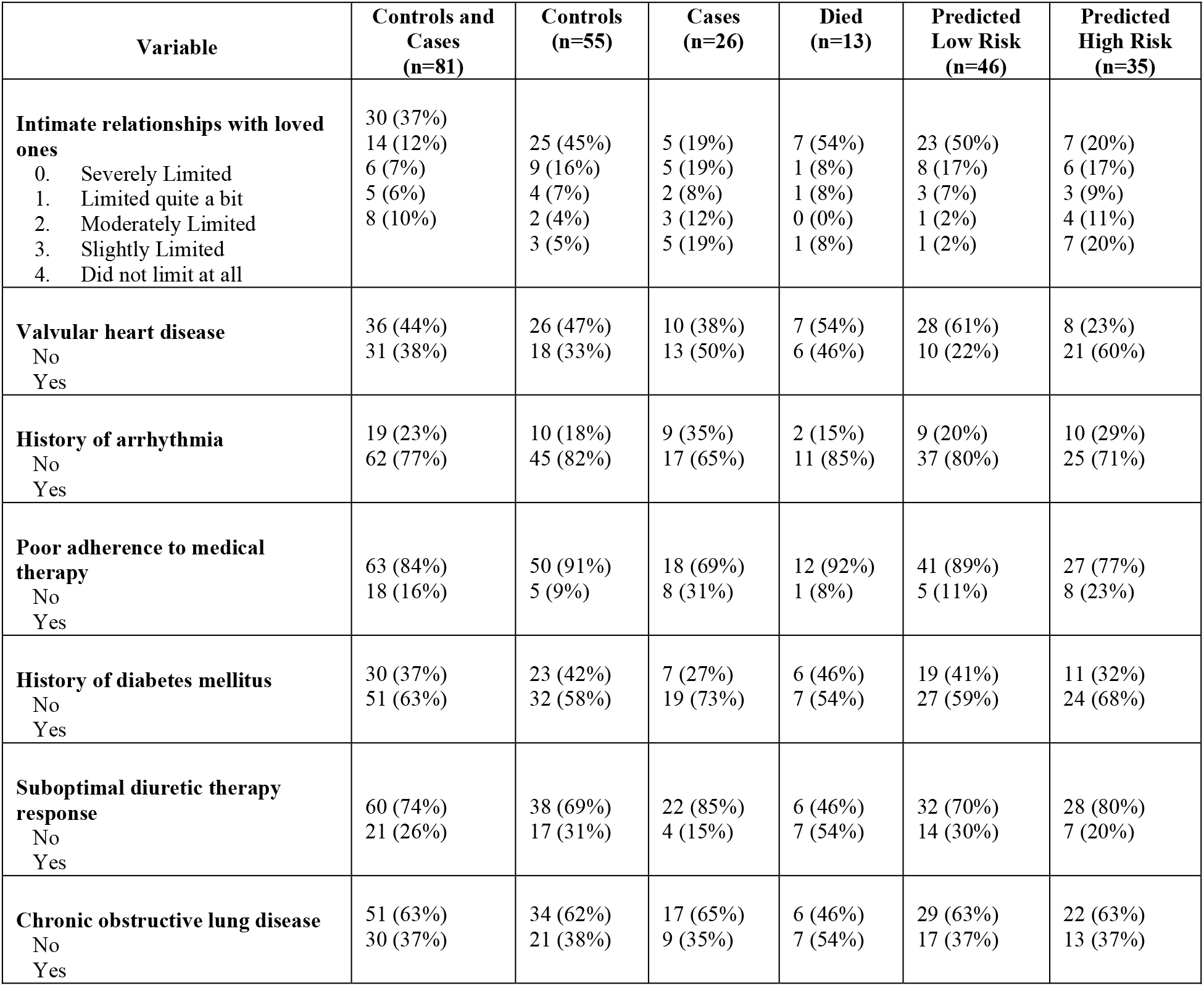
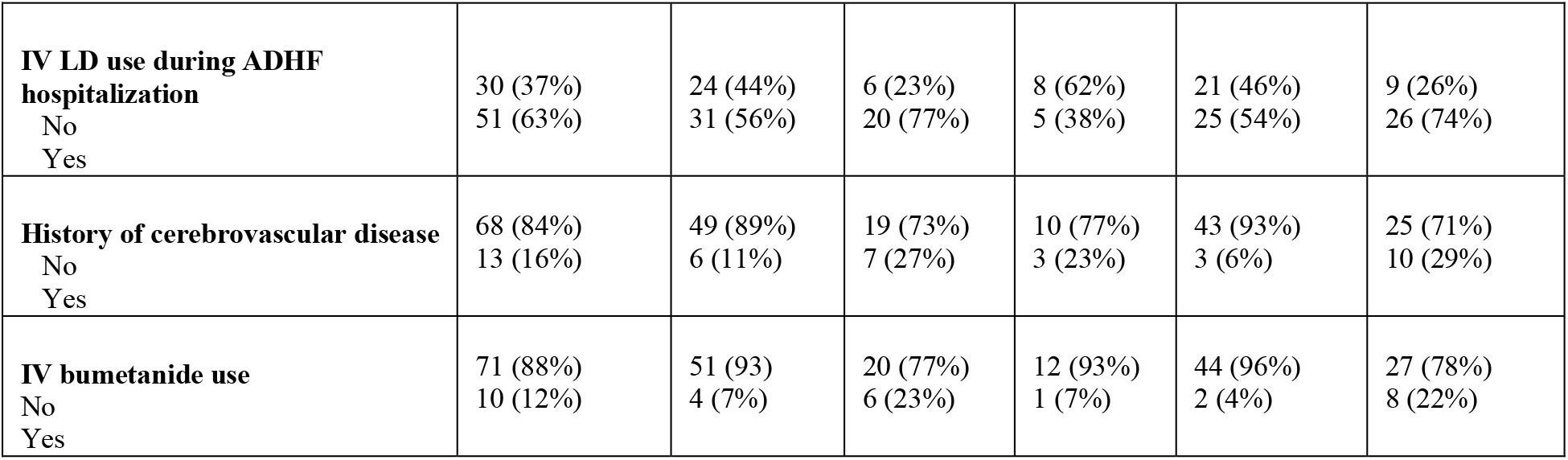
Characteristics of the variables of the final ensemble model. ADHF: acute decompensated heart failure; IV: intravenous; LD: Loop diuretic.

Sensitivity Analysis: We further implemented the proposed final ensemble model on 13 patients who died before the completion of a 90-day follow-up. Among these 13 patients, 7 did not have a 90-day event by our definition; 3 of those 7 were correctly classified by our model into no event. The other 6 of 13 did have 90-day HF events, and the proposed model predicted 2 of them as high risk. Finally, the proposed ensemble model predicted 46% of these 13 patients as high risk for HF events.

Table 6 summarizes the performance of our model when it was implemented on the IV LD arm (n=84). By comparing the model’s performance results in the Aquaphoresis (table 4) to the IV LD treatment group (table 6), it is evident that the proposed predictive model is designed to identify best responders to UF treatment rather than IV LD treatment. This could be attributed to different baseline characteristics that make them more responsive to UF than IV LD treatment.

**Table 6.** Confusion Matrix of LightGBM model for patients who received IV LD. IV: Intravenous; LD: Loop diuretic; NPV: negative predictive value; PPV: positive predictive value; SN: sensitivity; SP: specificity

| 90-day event |  | Actual 90-day event |  |  |
| --- | --- | --- | --- | --- |
|  |  | Yes | No |  |
| Predicted 90-day Event | Yes | 18 | 15 | PPV 55% |
|  | No | 22 | 29 | NPV 43% |
|  |  | SN 45% | SP 66% |  |

## 4. Discussion

Despite the advancement in diagnosis and management of heart failure, the 5-year mortality rate of HF adult patients is about 42%^22^. Reports have shown the total costs of HF were about $30 billion in 2012 and will be around $70 billion by 2030 (estimated increase cost rate of 127%)^23^. HF patients tend to require frequent hospitalization with high readmission rates compared to other diseases. Approximately 25% and 50% of HF patients are readmitted within 30 days and 6 months of discharge, respectively; 25% of these readmissions can be prevented^24^. Studies have shown that reducing HF readmission rates can reduce the cost of HF care hence the healthcare system burden and improve patient long-term survival and quality of life^25^.

HF clinical trials are costly, time-consuming and unguaranteed to result in positive results. Many factors can contribute to high trial failure rates, including patient selection, study design, and inability to monitor and follow-up with patients effectively during trials^26^. AI, with its different applications, can help to define appropriate patient phenotypes to avoid negative trials caused by factors other than the treatment(s) and support a study design of a lower sample size (refer to chart 1). In addition, AI will enable researchers to extract useful information from existing data and obtain very organized data input to design, conduct, and analyze more successful clinical trials. For instance, AI can avoid poor study design by finding similar studies addressing similar issues by performing a much more comprehensive literature review. Using AI and analyzing previous trials to determine the appropriate study size is another way to design a positive, successful trial^27^.

Many trials, including the RAPID-CHF, CARRESS-HF, and AVOID-HF trials, were designed to assess the role, safety, and efficacy of UF in HF. Those expensive and time-consuming trials were still inconclusive and lacked generalizability.

Using AI and ML, our study utilized existing data collected for the AVOID-HF trial to create a predicting model that can be used before initiating UF therapy to anticipate who will respond well to UF and who will be at high risk of future HF events. Our ML-designed model was more successful in predicting the outcome in patients with ADHF treated with Aquaphoresis. In the original AVOID-HF trial, the overall 90-day HF event rate in the UF arm was 32%, whereas our proposed predictive model was accurate in anticipating a 90-day HF event with much better statistical accuracy (AUC 0.88, SN 80%, SP 75%, NPV 90% and PPV 57%). In other words, if our model categorizes a patient as a super-respondent, the chance of a 90-day HF event is only 10%.

Besides its accuracy, our ML-model was able to examine the different variables and determine their significance. That led to identifying the top ten contributing factors that enhance the model’s applicability. For example, despite the daily use of the New York Heart Association (NYHA) functional classification of HF patients, the NYHA functional class was not a strong predictor of a 90-day HF event when compared to an intimate relationship with loved ones as was identified by our ML-model. This emphasizes on the advantage of AI and ML by “thinking outside the box” and finding unrecognized significant clinical factors. Our ML-designed predictive model can also be helpful in designing the next clinical trial for UF therapy in HF to avoid another neutral or negative trial caused by poor study design.

**Chart 1.**
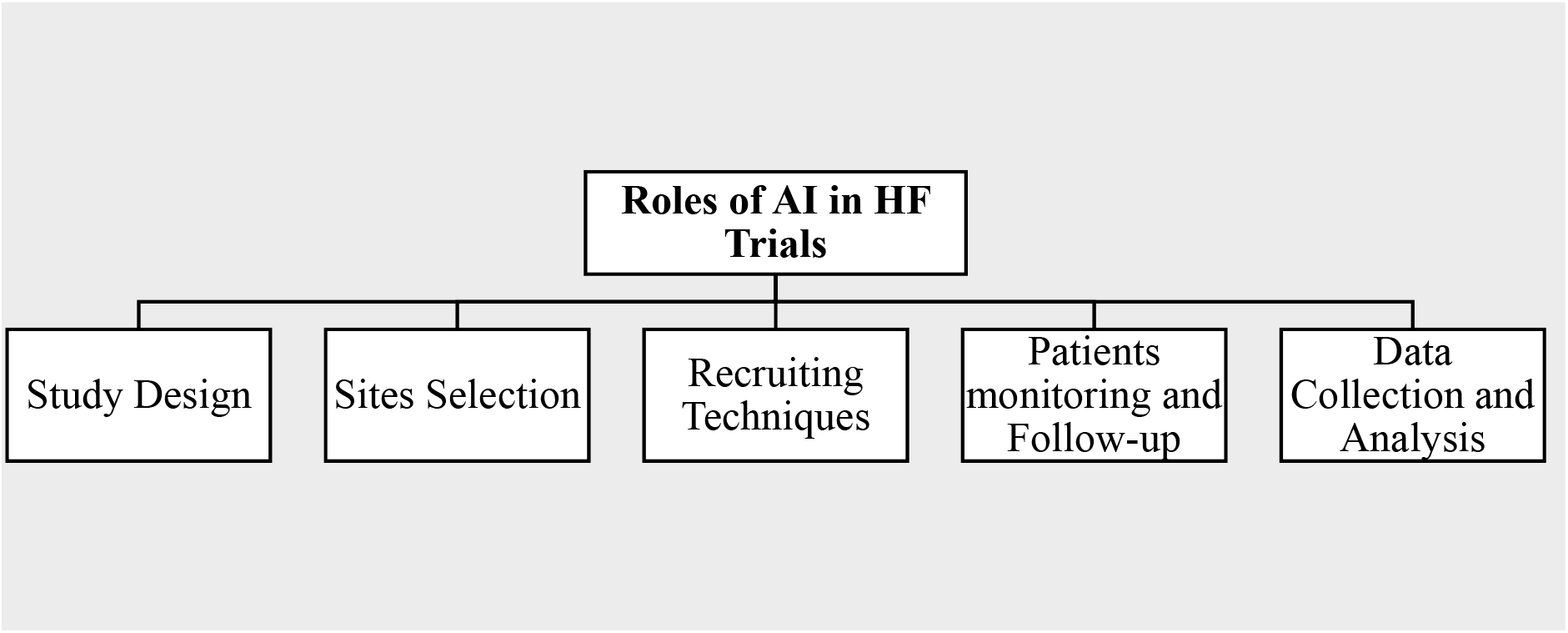
Roles of AI in HF trials. AI: Artificial intelligence; HF: Heart failure.

## 5. Limitations

Our ML-designed model is based on data from a single study. Our predictive model can benefit from an independent dataset for external validation, making it more generalizable. Despite its small sample size, we implemented a very comprehensive internal validation strategy by developing a leave-one-out cross-validation model. That led to good statistical outcomes (AUC, SN, SP, NPV, and PPV) which is by itself a predictor of successful external validation and generalizability.

## 6. Conclusions

Using existing data from the AVOID-HF trial, ML can better identify HF patients who will be responsive to Aquaphoresis therapy. This ML-predictive model was able to identify super-respondents to UF therapy and predict 90-day HF events better than the existing classic models. In addition, external validation of our model using an independent dataset will make it more generalizable to existing HF patients being considered for aquapheresis.

## Data Availability

Any requests for data should be made in writing to the corresponding author, and the data will be provided in accordance with relevant institutional and ethical guidelines.

## ABBREVIATIONS AND ACRONYMS

ADHF: acute decompensated heart failure
AI: artificial intelligence
AQ: Aquapheresis
HF: heart failure
IV: intravenous
LD: loop diuretic
ML: machine learning
UF: ultrafiltration

## PERSPECTIVES

### COMPTENCY IN PATIENT CARE AND PROCEDURAL SKILLS

Optimal use of UF in HF patients constantly evolves through clinical practice and trial learnings. AI and ML-based selection of an optimal patient for the application of UF can potentially lead to lower readmission rates.

### TRANSLATIONAL OUTLOOK

Due to the broad level of clinical experience in the utilization of UF in HF, the outcomes of such applications can lead to suboptimal clinical results, and this AI-based solution may help overcome these limitations.

## Notes

**AUTHOR DISCLOSURES:** The authors declare no conflict of interest.

### Competing Interest Statement

The authors have declared no competing interest.

### Funding Statement

The research received no external funding.

### Author Declarations

This study used data from the AVOID-HF (Aquapheresis versus Intravenous Diuretics and Hospitalization for Heart Failure) trial (all data was de-identified)

